# Unsupervised detection of SARS-CoV-2 mutations and lineages in Norwegian wastewater samples using long-read sequencing

**DOI:** 10.1101/2024.08.27.24312690

**Authors:** Ignacio Garcia, Rasmus K. Riis, Line V. Moen, Andreas Rohringer, Elisabeth H. Madslien, Karoline Bragstad

## Abstract

The COVID-19 pandemic has underscored the importance of virus surveillance in public health and wastewater-based epidemiology (WBE) has emerged as a non-invasive, cost-effective method for monitoring SARS-CoV-2 and its variants at the community level. Unfortunately, current variant surveillance methods depend heavily on genomic databases with data derived from clinical samples, which can become less sensitive and representative as clinical testing and sequencing efforts decline.

In this paper, we introduce HERCULES (<u>H</u>igh-throughput <u>E</u>pidemiological <u>R</u>econstruction and <u>C</u>lustering for <u>U</u>ncovering <u>L</u>ineages from <u>E</u>nvironmental <u>S</u>ARS-CoV-2), an unsupervised, database-independent method that uses long-read sequencing of a single 1 Kb fragment of the Spike gene. HERCULES identifies and quantifies mutations and lineages without requiring database-guided deconvolution, enhancing the detection of novel variants. We evaluated HERCULES on Norwegian wastewater samples collected from July 2022 to October 2023 as part of a national pilot on WBE of SARS-CoV-2. Strong correlations were observed between wastewater and clinical sample data in terms of prevalence of mutations and lineages. Furthermore, we found that SARS-CoV-2 trends in wastewater samples were identified one week earlier than in clinical data.

Our results demonstrate HERCULES’ capability to identify new lineages before their detection in clinical samples, providing early warnings of potential outbreaks. The methodology described in this paper is easily adaptable to other pathogens, offering a versatile tool for environmental surveillance of new emerging pathogens.

## Introduction

The COVID-19 pandemic had a significant impact on global health and has highlighted the effectiveness of virus surveillance as a driver of public health interventions to manage its transmission. Since effective surveillance of SARS-CoV-2 is crucial for identifying new variants and monitoring virus evolution, an unprecedented effort was dedicated to sequencing SARS-CoV-2 clinical samples during the peak of the pandemic. However, the end of the pandemic has brought a noticeable relaxation in these efforts^1^ (Fig. S1A). In that context, one surveillance approach that has recently gained attention is wastewater-based epidemiology (WBE), which has the potential to provide a fast, non-invasive, community-level snapshot of epidemiological situations using lower resources than the clinical surveillance and sequencing of clinical samples.

WBE involves searching wastewater for the presence of SARS-CoV-2 RNA, which can provide information on the presence of the pathogen in an area, the viral load and the prevalence of the different lineages of the virus in the population.

During the COVID-19 pandemic, several countries around the world implemented WBE to supplement traditional clinical surveillance methods. In Norway, a pilot project was initiated in June 2022 to test and evaluate the usefulness of wastewater-based surveillance of SARS-CoV-2^2^. While the evaluation of the pilot showed that wastewater surveillance provided early detection of new waves of infection, better analytical methods were needed to be able to assess the surveillance systems’ ability to detect new virus mutations and lineages^2^. With the increasing interest in WBE, a lot of effort has been put into developing new techniques to detect novel variants of SARS-CoV-2 and to monitor changes in their distribution over time, and, indeed, several WBE methods with short (Illumina) or long reads (Nanopore) have been published ^3,4,5^. While most of these methods use amplicon-based protocols with the viral genomes amplified in fragments^6,7^, the complexity of wastewater samples makes that sequencing methods tailored for clinical samples, such as tiled amplicon sequencing, might perform underperform in WBE analyses. While amplicon-based methods are very effective when all the viral particles in the sample are clones, they struggle to detect complex mixtures of viral strains and to identify co-occurring mutations since the different fragments of the viral genome are amplified with different efficiencies^8^. To cope with this limitation, most of the current methods to analyze SARS-CoV-2 lineages in wastewater rely on sequence databases obtained from clinical repositories such as GISAID^9^. Therefore, as soon as the sequencing efforts decline, the databases become incomplete, and new variants might remain undetected for a long time. This can cause these supervised database-based WBE analyses to become biased toward high-frequency lineages, and therefore their utility as surveillance methods to replace clinical-based sequencing is compromised. However, to the best of our knowledge, there is no state-of-the-art standard method for unsupervised analysis of SARS-CoV-2 sequencing data derived from wastewater samples.

In this paper, we describe HERCULES (*<u>H</u>igh-throughput <u>E</u>pidemiological <u>R</u>econstruction and <u>C</u>lustering for <u>U</u>ncovering <u>L</u>ineages from <u>E</u>nvironmental <u>S</u>ARS-CoV-2*), an unsupervised, database-free, method to identify and quantify SARS-CoV-2 mutations and lineages in wastewater samples. Our method is based on the amplification of one single small fragment which is then sequenced using long-read sequencing, thus, enabling the identification of genetic variations present on the same viral genome without the need for any database-guided lineage deconvolution^6^. Additionally, sequencing such a small fragment alleviates the sequencing costs and speeds up the surveillance, since the number of reads required to get enough depth to identify low-frequency lineages is approximately two orders of magnitude lower than the number of reads required by short read-based methods.

Despite these advantages, some challenges need to be considered, particularly when using long-read sequencing for WBE. One of the main challenges is the relatively high error rate of long-read sequencing^10,11^, which can lead to difficulties in the accurate identification of lineages, particularly when dealing with very low-abundance variants. To tackle this problem, we developed an algorithm to mask non-informative positions (i.e., positions that are identical in all the viral genomes of the sample) by identifying and ignoring positions with a noise value (i.e., the ratio of reads that do not agree with the most frequent base) consistent with the intrinsic error rate of long-read sequencing.

Moreover, we equipped HERCULES with tools to find and quantify lineages and to classify them according to the Pangolin nomenclature^12^. We have also included a *kmer* search tool to perform high-sensitivity searches of combinations of mutations or small sequences. To evaluate HERCULES, we analyzed wastewater samples acquired in Norway between July 2022 and October 2023 and we found a very good correlation between wastewater and clinical samples both in the mutation and lineage prevalences. We also discovered that the epidemiological trends of SARS-CoV-2 lineages seem to appear earlier in wastewater samples.

These results show the potential of HERCULES to identify novel emerging lineages before they have been assigned using any nomenclature and even before they have been detected in clinical samples. It can, therefore, be used to provide an early warning for potential outbreaks and help to guide public health interventions.

Moreover, although we created HERCULES with SARS-CoV-2 in mind, the methods and algorithms developed and implemented (i.e., noise identification, *mutation-lineage* calls, 2D-maps, etc.) are pathogen agnostic and can, therefore, be easily adapted to monitor other microorganisms in environmental samples.

## Results

### The incidence of Spike SARS-CoV-2 mutations is similar in wastewater and clinical samples

To evaluate the ability of HERCULES (Fig. 1A, Supplementary Methods) to find and quantify SARS-CoV-2 mutations and lineages in Norwegian wastewater, we regularly collected wastewater samples between July 26, 2022, and October 30, 2023, from five municipalities in Norway (Bergen, Oslo, Tromsø, Trondheim, and Ullensaker). We extracted the total RNA from the samples and amplified a 1 Kb DNA fragment from the SARS-CoV-2 Spike gene which was then sequenced using Oxford Nanopore Technology (ONT). The fragment that we selected starts at position 1250 of the Spike gene and overlaps with the Receptor Binding Domain (RBD), a part of the Spike protein involved in the binding of the virus to human cells. RBD is known for being very relevant for the fitness of the virus^13^ and, it is, indeed, one of the most mutated regions of the SARS-CoV-2 genome during the evolution of the virus (Fig. 1B).

**Figure 1:**
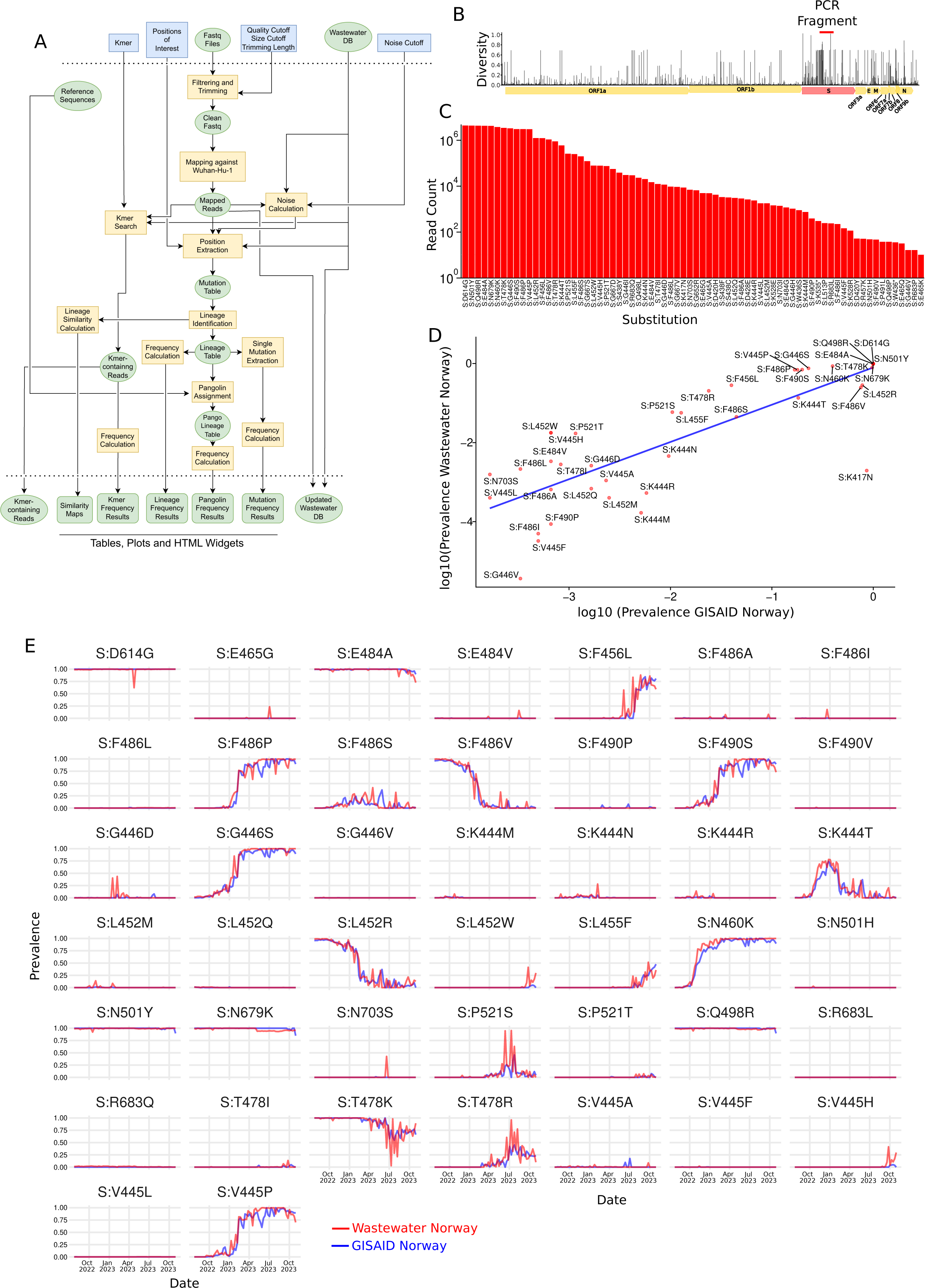
Identification of SARS-CoV-2 mutations in wastewater using HERCULES. **A.** HERCULES flowchart. **B.** Nextstrain’s^27^ output showing the nucleotide diversity in SARS-CoV-2 genome between 2020 and 2024. The red segment represents the genome fragment used in this paper. **C**. Bar plot showing the number of reads containing each of the mutations displayed on the X-axis. **D.** Scatter plot showing the correlation between the overall prevalence of the mutations identified in wastewater and the clinical samples from GISAID. The blue line represents the linear regression line between wastewater and GISAID prevalences. **E.** Line plots of the temporal prevalence of the mutations found both in wastewater (red lines) and clinical data from GISAID (blue lines).

In total, we collected 214 samples that were sequenced in 15 different batches. These sequencing results were further analyzed using HERCULES. Since the pipeline was designed to be a surveillance tool, we implemented a method to easily incorporate new data into a pre-existing analysis as a new batch is sequenced. In this paper, we present the latest analysis that includes all the samples.

We found that HERCULES could automatically identify and track 72 amino acid substitutions in the Spike protein (Fig. 1C). We analyzed whether these mutations had already been found in the clinical samples deposited in the GISAID database^9^, and we found that most of them had indeed been already reported in clinical samples (Table S1).

Then, we compared the prevalences of the mutations identified both in wastewater and clinical samples during the same period in Norway, and we found that both prevalences strongly correlate (Pearson’s correlation coefficient 0.77) and that overall, the correlation seems to be higher for high-prevalence mutations (Fig. 1D). This is not very surprising since the expected confidence intervals tend to be broader as the sample size decreases.

Interestingly, we noticed that 32 of the 72 mutations that we identified in wastewater samples were not present in the Norwegian clinical samples. These 32 cryptic mutations had low prevalences, ranging between 0.0002% and 2.8% in the wastewater (Table S1), and although most of them were eventually detected worldwide, their worldwide prevalences were even lower, ranging between 0.00004% and 0.22% (Table S1). Additionally, we spotted a mutation, S:K417N, with high frequency in clinical samples, but low frequency in wastewater (Fig. 1D). A deeper investigation of this discrepancy revealed that the nucleotide substitution responsible for the mutation S:K417N, overlaps with the forward primer (See Methods), and therefore it could never have been incorporated into any wastewater read.

We, therefore, masked S:K417N in all subsequent analyses. Although HERCULES was designed to identify new mutations independently, it also contains an internal, but updatable, database of viruses, and it uses this database to inform on possible mutation sites to track (See Supplementary Methods for details). We found that the S:K417N mutation was tracked because of this pre-processing step. These results suggests that although some of the cryptic mutations could be the result of sequencing errors, others might have been never detected in clinical samples in Norway because of their very low abundance in the population.

Next, we explored the temporal prevalences of the mutations identified in wastewater, and we compared them with their prevalences in clinical samples. As expected, we found that different mutations had different prevalence patterns (i.e., increasing, decreasing, stable, increasing-decreasing) (Fig. 1E, red lines). After analyzing the prevalence of the same mutations in the Norwegian clinical samples (Fig. 1E, blue lines), we found a very strong overlap between the prevalences of the mutations in clinical samples and wastewater (Fig. 1E).

These findings support the use of unsupervised methods on wastewater samples to estimate the epidemiological trends of SARS-CoV-2 mutations in the clinic.

### HERCULES can be used to identify “low-level” SARS-CoV-2 lineages

To evaluate how the mutations tracked were combined into different viral lineages, we tracked the unique combinations of mutations that appeared together in the same read. We named these unique combinations of mutations *mutation-lineages* and they represent the lowest possible level to classify viral lineages. We found that although HERCULES identified 487 unique *mutation-lineages,* only 29 of them reached a prevalence higher than 4% and appeared in at least 10% of the samples. We labeled these high-frequency *mutation-lineages* as <u>N</u>orwegian <u>W</u>astewater <u>L</u>ineages 1 to 29 (Fig. 2A, red lines).

**Figure 2:**
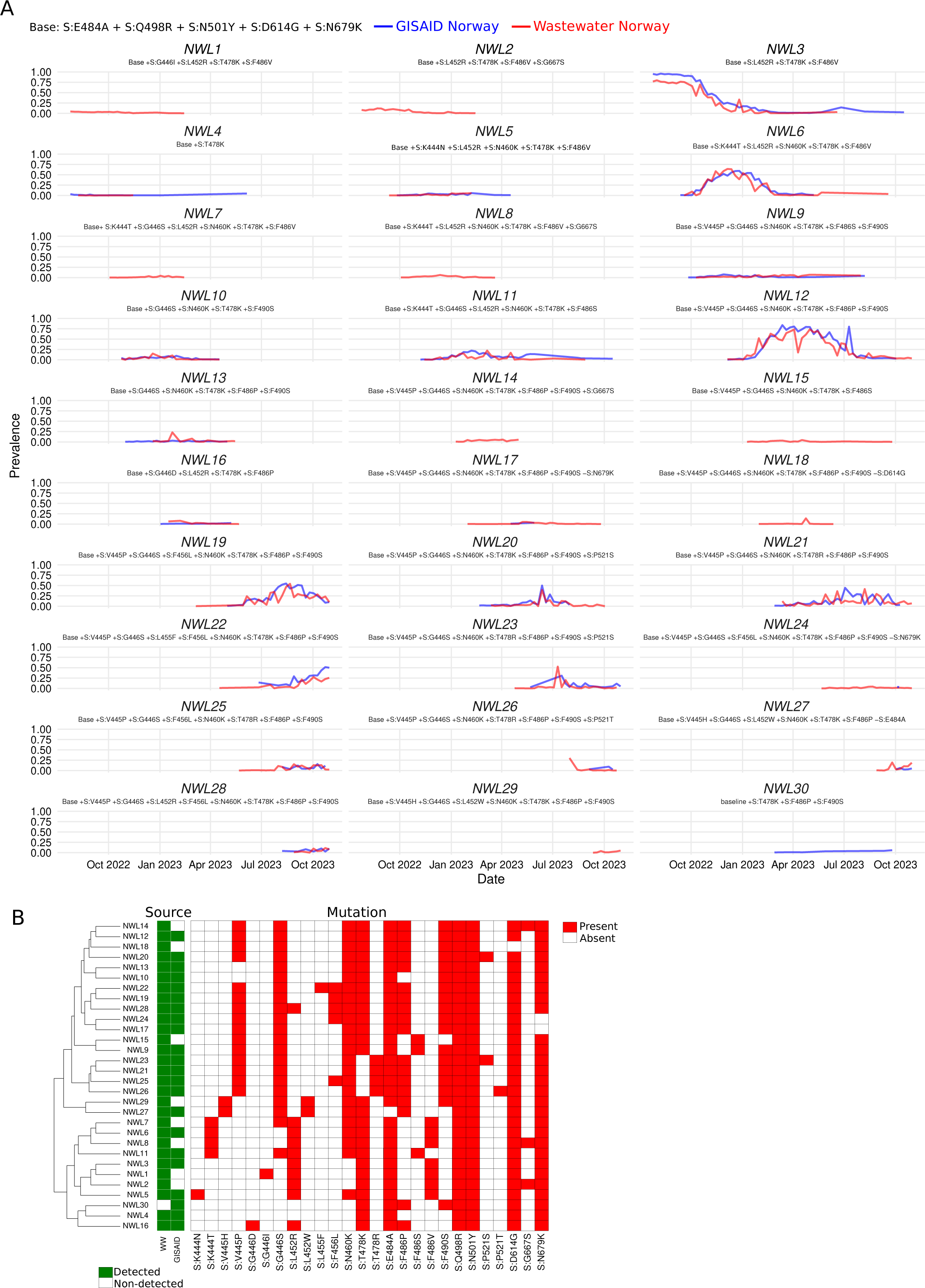
Identification of SARS-CoV-2 “low level” lineages in wastewater using HERCULES. **A.** Line plots of the temporal prevalence of the *mutation-lineages* (i.e., the different combinations of mutations present in the same read) present in wastewater (red lines) and clinical data from GISAID (blue lines). The mutations present in each of the *mutation-lineages* are displayed. The different *mutation-lineages* were labeled as <u>N</u>orwegian <u>W</u>astewater <u>L</u>ineages. **B.** Tile plot and dendrogram showing the mutations present in each *mutation-lineage* and the genetic distance between the lineages. The mutations present in each lineage are shown as red-filled tiles.

To determine whether there was an overlap between the lineages flagged by HERCULES and the clinical samples, we downloaded from GISAID the 8087 Norwegian sequences within the same time window. We aligned them to the Wuhan-Hu-1 reference, extracted the region of interest, and saved them in a HERCULES-compatible format (See Methods for details). Then, we ran HERCULES on them to identify the *mutation-lineages* present in the clinical samples (Fig. 2A, blue lines). By analyzing the clinical samples, we identified an additional lineage, NWL30, that was only found on GISAID and we discovered that eight *mutation-lineages* (NWL1, NWL2, NWL7, NWL8, NWL14, NWL15, NWL18, and NWL29) were wastewater-specific lineages (Fig. 2A and 2B).

As before, we found that there is a very good correlation between the average incidence (Fig. S1B) and the temporal patterns of the lineages found both in wastewater and clinical samples (Fig. 2A).

Prompted by the presence of cryptic *mutation-lineages* (i.e., *mutation-lineages* that were found in wastewater but not in clinical samples), we decided to systematically investigate whether they could be the result of sequencing errors or artifacts derived from the analysis. After a deeper analysis of the cryptic *mutation-lineages*, we found that (i) NWL1 and NWL2 were very similar to NWL3. Indeed, NWL1 is NWL3 plus the S:G446I mutation and NWL2 is NWL3 plus S:G667S (Fig. 2B). (ii) NWL7 and NWL8 are very similar to NWL6. NWL7 is NWL6 plus S:G446S and NWL8 is NWL6 plus S:G667S (Fig. 2B). (iii) NWL14 is NWL12 plus S:G667S. (iv) NWL15 is similar to NWL9, but without S:F490S (Fig. 2B). (v) NWL18 is NWL12 without S:D614G (Fig. 2B), and finally (vi) NWL29 is NWL27 plus S:E484A and S:F490S (Fig. 2B).

We noticed that there was one common mutation (i.e., S:G667S) accounting for the differences between three wastewater-specific lineages (NWL2, NWL8, and NWL14) and their closest non-cryptic counterparts (NWL3, NWL6 and NWL12 respectively). Interestingly, S:G667S has never been detected in clinical samples in Norway. After extracting more than 150K reads from the lineages NWL2, NWL8 and NWL14, we found that all of them carried the substitution that drives the S:G667S mutation (i.e., G23561A). Moreover, most of the S:G667-containing reads do not contain deletions in the surroundings of the 667^th^ codon (i.e., plus/minus five nucleotides) that could explain the presence of the substitution.

Therefore, due to the large number of reads carrying it, we concluded that the S:G667S mutation was most likely present in wastewater samples. It is possible that the mutation has never been observed in clinical samples because of the lower sensitivity of clinical sampling. Similarly, we analyzed the S:G446I-positive sequences since S:G446I is the mutation that accounts for the divergence between NWL1 and NWL3. Again, we found that it has never been detected in Norwegian clinical samples. After analyzing all the reads assigned to NWL1 (N=30825) we found that 100% of them carried the mutations G22898A and G22899T. The effect of both mutations results in the S:G446I substitution. It is likely that the S:G446I mutation occurred as a subsequent mutation on a lineage carrying the G22898A mutation that causes the S:G446S substitution, a mutation already present in several omicron lineages. Again, due to the large number of reads supporting it and the absence of deletions around the 446^th^ codon, we concluded that the presence of the mutation S:G446I in wastewater samples does not seem to be an artifact. We also confirmed that the lineage NWL7 (N=21529 reads) actually contains the S:G446S substitution encoded by the mutation G22898A, that the lineage NWL29 (N=13874 reads) carries S:F490S and S:E484A and that the reads of the lineage NWL15 (N=26915 reads) do not have any nucleotide substitution that could account for the S:F490S mutation.

Lastly, we checked whether the NWL18 lineage carried the mutation S:D614G, a mutation acquired very early in the pandemic^14,15^. We found that although none of the reads (N=13731) carried the S:D614G mutation (A23403G), the lineage was heterogeneous regarding the 614^th^ codon: 10.9% of the reads seemed to have the reversion S:G614D and 84.5% of the reads had at least one deletion mapped into the codon. By exploring the surroundings of the 614^th^ codon we found that indeed a very significant fraction of reads (89.4%) had at least one deletion in the area (Fig. S1C). In contrast, other lineages (e.g., NWL15) have the A23403G mutations in 99.7% of the reads and no deletions are present in the area (Fig. S1C). We, therefore, conclude that although it seems that the absence of S:D614G might be artefactual for a very significant fraction of reads, some viruses of the *mutation-lineage* might have reverted the S:D614G substitution. Unfortunately, false positive deletions are very common in ONT reads, and therefore it is very difficult to confirm analytically if all the deletions are real and if so, which amino acid changes these deletions might cause.

As in the case of the cryptic mutations, the incidences of the cryptic *mutation-lineages* are low in wastewater (Fig. 2A and Fig. S1B), suggesting that the lower sensitivity of clinical sampling could be the reason for their absence in clinical samples.

Except for the absence of S:D614G in NWL18, we could not find evidence suggesting that the observed results were due to any artifact, thus validating the use of unsupervised methods to find and track emerging SARS-CoV-2 lineages in wastewater. The results also show that intensive wastewater analysis can be used to uncover cryptic mutations and lineages.

Moreover, the good correlation that we have observed between clinical and wastewater sources suggests that the SARS-CoV-2 surveillance strategy implemented in Norway during the 2022-2023 session was reasonable, unbiased, and representative.

The prevalences of “high-level” SARS-CoV-2 lineages in the population can be effectively estimated from a partial Spike fragment from wastewater.

Since very early in the pandemic, the Pangolin nomenclature has been the *de facto* method to classify SARS-CoV-2 lineages because of its robustness and dynamic nature^12,16^. However, the increasing number of concurrent genetically similar Pangolin lineages makes it difficult to draw efficient epidemiological assessments using Pangolin nomenclature. For example, 596 different Pangolin lineages have been detected among the 8087 Norwegian sequences deposited in GISAID during the study period (Fig. S1D). However, only 6.5% of them have a maximum incidence larger than 10% (Fig. S1D). To mitigate the effects of this very high granularity, several methods to aggregate Pangolin lineages are broadly used in SARS-CoV-2 surveillance such as the “high-level” nomenclature proposed by the World Health Organization (WHO) (i.e., Alpha, Beta, Delta, etc.)^17^, or the classification method proposed by The European Surveillance System (TESSy)^18^ (ECDC, 2024) (Table S2).

To make HERCULES’ results compatible with other available epidemiological sources, we implemented a method to classify *mutation-lineages* as Pangolin lineages that could be further aggregated into higher-level lineages such as those defined by TESSy or WHO. Since we are only using a small fraction of the genome and many Pangolin lineages differ from each other in mutations outside the sequenced region, it is expected that the Pangolin assignment would eventually produce ambiguous results. To minimize this ambiguity, HERCULES uses the phylogenetic information encoded in the Pangolin nomenclature to compress the Pangolin lineages matching them to the closest common parental lineage. For example, although the sequences EPI_ISL_14773382 and EPI_ISL_15168744 contain the same mutations in the sequenced region (i.e., G22813T, T22882G, T22917G, G22992A, C22995A, A23013C, T23018G, A23055G, A23063T, T23075C, A23403G, C23525T, G23587T, T23599G and C23604A), they are classified as BA.5.2.9 and BA.5.2.1, respectively. In this case, HERCULES would classify them both as BA.5.2.X.

To evaluate the efficiency of HERCULES in the “high-level” lineage assignment task, we decided to use the TESSy classification method since it provides a medium granularity and a good dynamic range in the time window considered (Fig. S1E).

We found that after the Pangolin assignment and the aggregation into TESSy’s lineages, the wastewater results matched the results found in the clinic for the high incidence TESSy lineages: BA.5, BA.2.75, XBB.1.5, XBB.1.5-like+F456L, BQ.1 and BA.2.86 (Fig. 3A) suggesting that HERCULES performed fairly well classifying reads into “high-level” lineages. Unfortunately, HERCULES could not resolve the TESSy lineage for reads assigned to some unresolved Pangolin lineages (i.e., Pangolin lineages BA.X, BA.2.X, XBB.1.5.X, XBB.X and B.1.X) (Fig. 3B).

**Figure 3:**
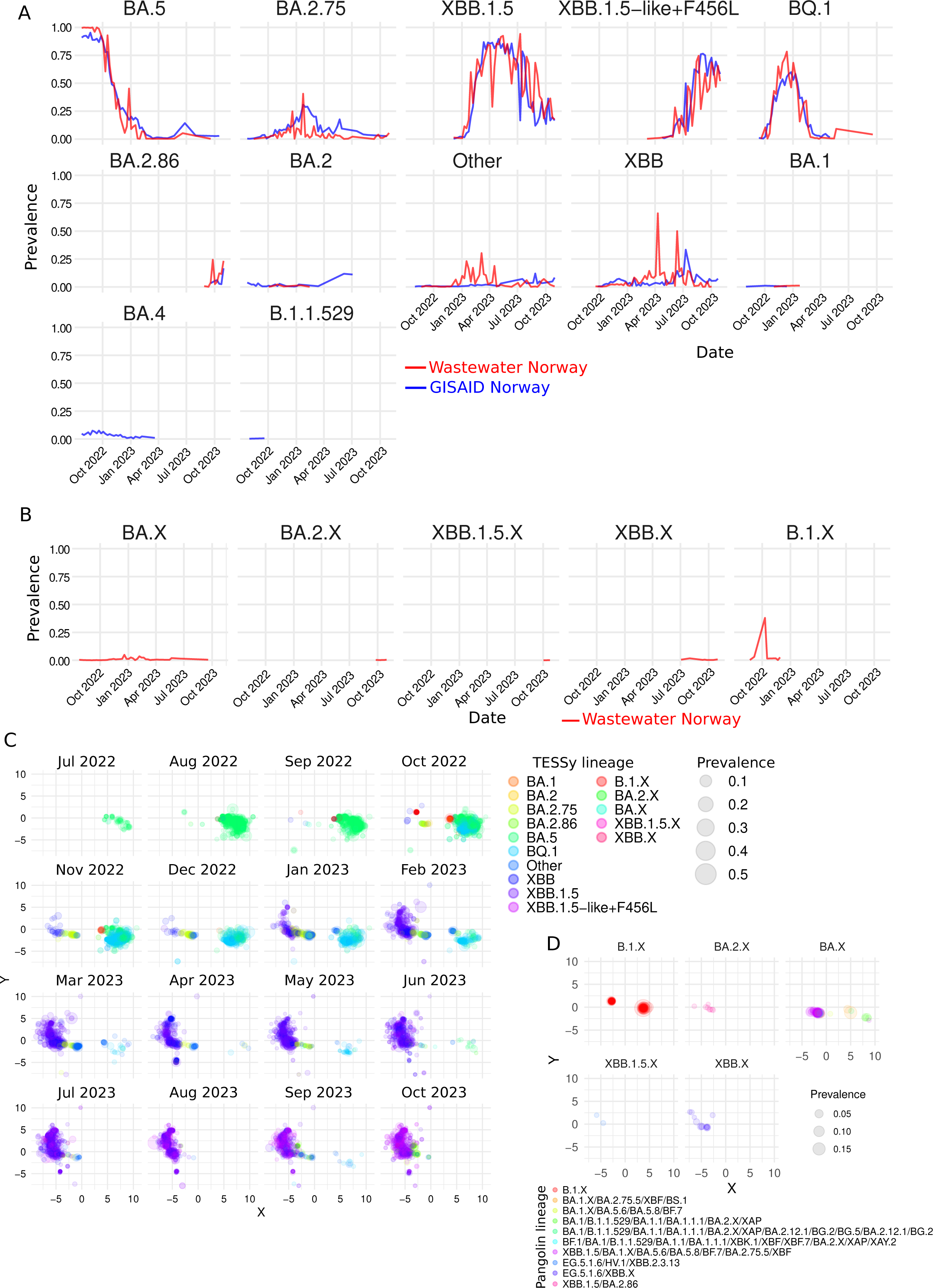
Identification of SARS-CoV-2 TESSy lineages in wastewater using HERCULES. **A.** Line plots of the temporal prevalence of The European Surveillance System (TESSy) lineages present in wastewater (red lines) and clinical data from GISAID (blue lines). **B.** Line plots of the temporal prevalence of lineages that could not be assigned to any TESSy lineage. The TESSy lineages were compressed to the closest common lineage (See Methods for details). **C.** Temporal 2D maps of the *mutation-lineages* present in the samples during each month. The coordinates to represent each point in the map were obtained from compressing the probabilities of each *mutation-lineages* to belong to different Pangolin lineages. Closer points are genetically closer (See Methods). The different colors represent the different TESSy lineages and the sizes of the points reflect their prevalences. **D.** 2D maps of the *mutation-lineages* present in the samples that could not be assigned to any TESSy lineage (Fig. 2B). The different colors represent the different Pangolin Lineages that the *mutation-lineages* were assigned to, and the size of the points their prevalences.

To further investigate these unresolved lineages and to provide a deeper overview of the heterogeneity of the samples in a time-wise manner, we developed a probabilistic approach to map *mutation-lineages* into a 2D plot (See Supplementary Methods for details) (Fig. 3C). This method uses the probability of a read belonging to the lineages present on a predefined, but updatable, internal database to plot similar sequences closer in the lineage’s map. This probabilistic map can be used to identify novel lineages, outliers, and even contaminations in the samples. HERCULES outputs these results as static plots, tables, and dynamic HTML widgets which can be visually inspected to find out the Pangolin lineage assigned, the mutations present, the discrepant mutations according to the Pangolin lineage assigned to, and the incidence of all *mutation-lineages* present in the samples (Fig. S1F). Although this 2D-mapping method depends on an internal database, novel unknown lineages will be placed separately from other different lineages on the map.

By exploring the 2D-maps, we noticed that although the sequences compressed into the B.1.X lineage (Fig. 3B) belonged to two distinct groups (Fig. 3C and Fig. 3D), HERCULES classified them both as Pangolin B.1.X. Interestingly, we noticed that the two B.1.X groups had very low dispersion in the map, suggesting few differences among them at the mutation level. After investigating the clean mutations (i.e., mutations not caused by spurious frameshifts due to sequencing errors) present in the unresolved lineage B.1.X, we found that indeed 98.6% of the reads had less than three mutations, 54.1% of them carried just the S:D614G and 44.5% of the reads had different combinations of S:D614G and just another mutation. The reduced number of mutations, together with the fact that the lineage peaked in October 2022 in just one location (i.e., Bergen), make us hypothesize that the B.1.X lineage could be the result of the contamination of a sample with the positive control that was included in the experiment alongside the wastewater samples since the positive control (GISAID ID: EPI_ISL_449791) has just one mutation (S:D614G) on the Spike gene.

Next, we analyzed the unresolved BA.2.X lineage and we found that HERCULES could not resolve some reads between Pangolin XBB.1.5 and BA.2.86, and therefore it assigned them all to the closest common Pangolin (i.e., BA.2.X). By analyzing the mutations carried by the BA.2.X reads, we found that all of them carried the BA.2.86 specific mutation S:L452W, but none of them had the mutation S:V445H, which is another BA.2.86 specific mutation.

Interestingly, among the Norwegian clinical samples obtained in 2024, 4.5% of the S:L452W-carrying viruses do not possess S:V445H. Interestingly, the first time that a S:L452W-virus lacking S:V445H was detected in a clinical sample was the 2^nd^ of January 2024 while the lineage BA.2.X was constantly detected in wastewater samples since the 25th of September 2023, suggesting that the S:L452W-positve/S:V445H-negative lineage was actually present in the population for a long time before it was detected on any clinical sample.

Next, we found that all the reads assigned to the Pangolin XBB.1.5.X lineage could not be resolved between the Pangolin lineages EG.5.1.6, HV.1, and XBB.2.3.13 (Fig. 3D) and that all the XBB.X came from reads that could not be resolved between EG.5.1.6 and XBB.X (Fig. 3D).

Finally, we found that the group of reads assigned to the BA.X fell into six different categories of unresolved Pangolin lineages (Fig. 3D).

Considering the very low incidence of the unresolved lineages, it is difficult to further assess if they could not be resolved because of sequencing errors introduced on the reads or if they could not be resolved by the limitation of analyzing just a a small fragment of the genome. In either case, HERCULES performed quite well assigning *mutation-lineages* to Pangolin lineages that could be further aggregated into higher-level lineages. Moreover, the TESSy lineages detected seem to be fully consistent with the mutations carried by the reads that they carry.

### High-sensitivity search methods show that the mutation trends observed in the clinic appear earlier in wastewater samples

To find *lineage-defining* mutations that might have not reached the threshold to be tracked by the noise-detection algorithm, we equipped HERCULES with a *kmer* search method (See Supplementary Methods for details) thus allowing the identification and quantification of different combinations of point mutations or small DNA sequences in the wastewater samples. This *kmer* search is the method with the highest possible sensitivity given the systematic uncertainty of the ONT.

To test this approach, we screened wastewater samples searching for all the nucleotide substitutions detected in clinical data (Fig. 4A and Fig. 4B). In total, we tracked 390 nucleotide substitutions that accounted for 234 mutations at the amino acid level. Next, we computed the differences in their weekly prevalences between wastewater and clinical data (Fig. 4C). Although we found that the discrepancies were very low, we noticed that they tend to be higher after July 2023 (Fig. 4C and Fig. S1G), which is when the sequencing effort in Norway fell under 50 sequences per week (Fig. S1H). This is not surprising, as the uncertainty in prevalence naturally increases when the sample size decreases.

**Figure 4:**
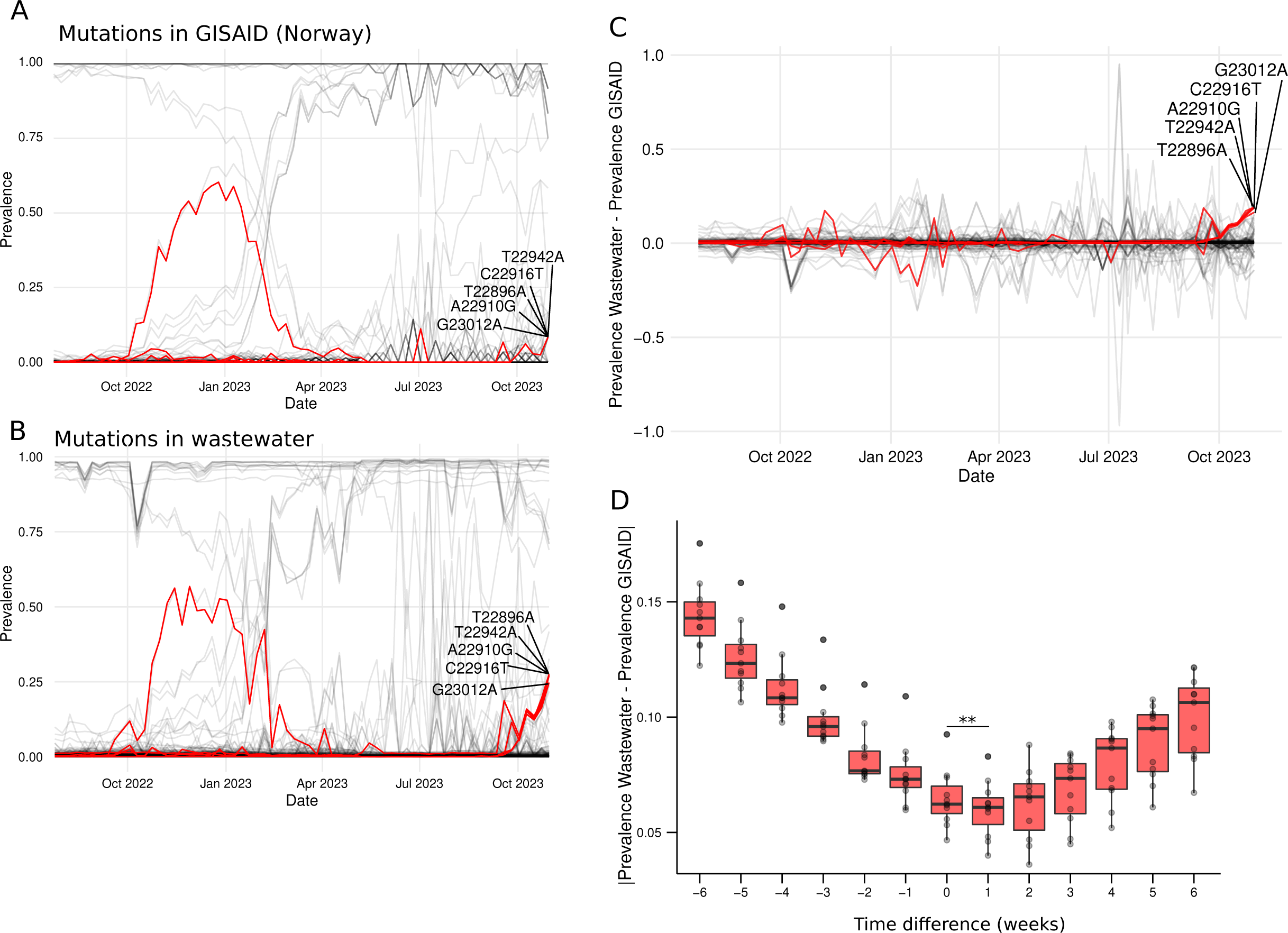
Identification of point mutations using high-sensitive methods and detection timing evaluation. **A.** Line plot of the temporal prevalence of nucleotide mutations identified in clinical samples obtained from GISAID. Each line represents a nucleotide mutation. The prevalences of T22896A, A22910G, T22942A, C22916T, and G23012A are shown with red lines. **B.** Line plot of the temporal prevalence of nucleotide mutations identified in wastewater samples. Each line represents a nucleotide mutation. The prevalences of T22896A, A22910G, T22942A, C22916T, and G23012A are shown with red lines. **C.** Line plot of the differences in prevalence between wastewater and clinical samples (GISAID). Each line represents a nucleotide mutation and on the Y-axis the differences in their ratios are shown. The difference in the prevalences of T22896A, A22910G, T22942A, C22916T, and G23012A are shown with red lines. **D.** Boxplots of the absolute differences i n prevalence between wastewater and clinical samples (GISAID) for the 34 mutations with GISAD prevalences ranging between 40% and 60% from January to April 2023 (Fig. 4B) when the collection times of the wastewater samples are moved backward and forward in time. On the X-axis the time displacement applied to the wastewater samples is shown. ** represents a p-value < 0.01.

After a closer look at the data, we found a set of five mutations (T22896A, A22910G, T22942A, C22916T, and G23012A) with overlapping prevalences that seem to increase in wastewater samples after October 2023, while their frequencies remained low in the clinical samples of the same period (Fig. 4A, Fig. 4B and Fig. 4C).

A deeper analysis of these mutations showed that their increase was concomitant with the rise of BA.2.86 derived lineages. Indeed, the five mutations are very prevalent in BA.2.86 lineages and they became highly dominant, being present in more than 87% of the Norwegian clinical samples sequenced in 2024.

This finding suggests that the incidence trends might appear earlier in wastewater samples than in clinical samples as it was also suggested by the early detection of the *mutation-lineage* carrying S:L452W but not S:V445H. To test this hypothesis, we selected the 34 mutations with prevalences between 40% and 60% from January to April 2023. By selecting mutations within that prevalence range and time window, we ignored the mutations with little changes in their prevalence because they could bias the analysis.

Next, we computed how the differences in the prevalences between wastewater and GISAID changed as we artificially displaced the wastewater sampling dates back and forth (Fig. 4D). Interestingly, we found that the differences between wastewater and clinical samples were minimal when we displaced the wastewater sampling dates one week forward (i.e., we pretended that the wastewater samples were taken one week later than their actual sampling week) (Fig. 4D).

These results confirm the hypothesis that the changes in prevalence can be detected in wastewater before they appear during clinical surveillance, and it sets this advanced detection window to one week.

Altogether, we have found a very good correlation between clinical samples and wastewater in all the analyses included in HERCULES, and it shows that using long-read sequencing of a small Spike fragment can be used to effectively estimate the prevalence of SARS-CoV-2 mutations and lineages at the population level.

## Discussion

WBE methods have recently gained substantial interest as pathogen surveillance tools since they can provide a good overview of the prevalence of a pathogen in the population^19,20^ However, most of the WBE approaches to quantify pathogen lineages rely on previous knowledge of the lineages to be found^6,7^. This weakness makes them unable to identify and track new emerging lineages. Moreover, the necessity of updated databases makes these methods highly dependent on clinical sampling and, therefore, not suited to replace, the sequencing of clinical samples during surveillance.

In this paper, we present HERCULES, an unsupervised, database-independent WBE method that uses long reads on a small fragment of the genome to estimate the prevalence of different mutations and lineages. Combining long-read sequencing of a small fragment of the genome with unsupervised analysis has several advantages: (i) the pipeline does not require a dataset to identify and track novel lineages with previously unseen mutations. (ii) There is no need for lineage deconvolution^6^; therefore, the theoretical sensitive limit is much lower, especially when using *kmer* searches. This point becomes very relevant in the case of SARS-CoV-2 whose genomes seem to be highly fragmented in wastewater samples^21^. (iii) Since the number of reads required to get the same coverage is much lower than when using tiled amplicon approaches, the sequencing cost per sample and the sequencing times are much lower. This would make it possible to include more sampling points without increasing the costs. Moreover, cheaper approaches would allow low-income countries to implement better WBE methods, making worldwide surveillance more comprehensive. We used HERCULES to analyze the presence of different SARS-CoV-2 mutations and lineages in Norway during 2022-2023. Using HERCULES, we were able to find relevant SARS-CoV-2 mutations and to quantify their prevalences over time. We found that the weekly prevalences of these mutations were very similar between wastewater and clinical samples.

Moreover, HERCULES managed to track how the different mutations were combined to form *mutation-lineages* and accurately quantify their prevalences. We also found a very good correlation between the *mutation-lineages* found in the clinic and wastewater. In addition, we found that HERCULES managed to correctly classify most of these *mutation-lineages* into Pangolin lineages, making HERCULES’ results easy to integrate with other Pangolin-based epidemiological sources.

One interesting feature of wastewater analyses is that they can be used to detect cryptic lineages^22^, which can provide important insights into the epidemiology and evolution of a pathogen. Interestingly, we discovered some cryptic SARS-CoV-2 lineages in wastewater. However, their low prevalences suggest that they were not detected in the clinic probably due to the reduced sensitivity of clinical sampling and not because of differences in the biology of the virus (e.g., lineages or mutations causing milder symptoms would theoretically appear in wastewater much more frequently than in the clinic).

Lastly, we studied the timing at which epidemiological trends occur in wastewater when compared with the clinic, and we found that the epidemiological patterns seem to appear in wastewater one week before they are found in the clinic. Although other studies have found equivalent results by comparing the dates of first time that a lineage in found in the clinic and in wastewater^6^; in this study, we used the entire prevalence trend of 34 mutations. By analyzing the prevalence of several mutations during a long period, the bias due to the lower sensitivity to spot the first occurrence of a mutation or lineage disappears. In any case, this finding makes WBE a valuable tool to forecast epidemiological trends, especially when combined with epidemiological modeling.

Despite the advantages of using HERCULES in WBE analyses, some limitations must be considered when using it for surveillance purposes. The main limitation comes from the use of a small fragment of the genome to classify viruses into lineages. This causes unsolvable uncertainties when classifying lineages that only diverge in mutations outside the sequenced Spike region. Although these uncertain lineages represent a small fraction in this study and their effect on the aggregation of lineages is very limited, this might be different for future lineages.

This limitation could be minimized by using larger genomic fragments. Indeed, HERCULES can currently handle larger Spike fragments without any adjustment (See Supplementary Methods).

Another limitation of HERCULES is the relatively high error rate of long-read sequencing causing the introduction of false-positive mutations into the analyses. This problem can be tackled by imposing different cutoffs to call mutations into the analyses. In this study, we have used low-stringency cutoffs to be able to evaluate the sensitivity of the method, and indeed we have detected a false-positive mutation (i.e., S:K417N). Finding the optimal cutoff must be evaluated for each use case since it will result in differences in the sensitivity and specificity of the method. The high error rate is particularly problematic when it causes the call of false insertions and deletions onto the reads. To handle the presence of false indels, HERCULES partially ignores the presence of frameshifts due to deletions in some of its outputs and it just totally ignores any insertion on the reads.

In conclusion, monitoring wastewater is a promising tool for early detection and surveillance of SARS-CoV-2 lineages and in this paper, we have shown that unsupervised approaches such as HERCULES can be used to replace, at least partially, sequencing clinical samples to monitor SARS-CoV-2.

To facilitate the use of this method, we have released HERCULES as a *Docker-based ready-to-use* pipeline that integrates all required tools (mutation finder, lineage mapper, *kmer* searcher, etc.) to analyze SARS-CoV-2 mutations and lineages in wastewater samples. Although HERCULES was designed to track SARS-CoV-2 lineages, it is possible to easily adapt the pipeline to track lineages of any other pathogen of interest or even to perform other metagenomic analyses when databases are sparse or missing.

## Methods

### Wastewater sampling, collection and preparation

Samples were collected through the Norwegian pilot on wastewater-based surveillance of SARS-CoV-2 running in the period June 2022-November 2023^2^. Untreated wastewater samples were collected weekly or bi-weekly from the inlet of wastewater treatment plants receiving wastewater form the largest urban municipalities in Norway. Samples were either collected through 24–72 hour flow-proportional composite sampling, time-proportional composite sampling, or grab sampling. The samples were then transferred into 1-liter bottles and kept at 4°C, before being shipped on ice to the contract laboratory (Nemko Norlab) within 24 hours. At the laboratory, total nucleic acid was extracted and analyzed for the detection of SARS-CoV-2 RNA using RT-qPCR. The sample extracts from SARS-CoV-2 positive samples were pooled into mixed samples, each pool consisting of total nucleic acid extracts from the same municipality sampled within the same calendar week and stored frozen at -16°C before shipped to the sequencing reference laboratory at the Norwegian Institute of Public Health (NIPH) where they were stored at -80°C before sequencing.

### PCR amplification and library preparation

A 1049 bp nucleotide fragment was amplified with PCR as described by Rasmussen et al.,^23^. A PCR mix consisting on 10 µl OneStep Primescript mix, 0,5 µl Superscript IV RT/PT HF enzyme mix, 0,4 µM of each primer (i.e., ARTIC V3 (https://artic.network/ncov-2019) primers *nCoV-2019_76_LEFT_alt3* (5’-GGGCAAACTGGAAAGATTGCTGA-3’) and *nCoV-2019_78_RIGHT* (5’-TGTGTACAAAAACTGCCATATTGCA-3’)), and 1 µl diluted total nucleic acid extracts from wastewater samples.

The amplified of a 1049-nucleotide fragment purified with Agencourt AMPure XP Beads before a second PCR for Nanopore barcoding with t*ailed-76_LEFT_alt3* (5’-TTTCTGTTGGTGCTGATATTGCGGGCAAACTGGAAAGATTGCTGA-3’) and *tailed-78_RIGHT* primers (5’-ACTTGCCTGTCGCTCTATCTTCTGTGTACAAAAACT GCCATATTGCA-3’). Then, a third PCR was used to attach the Barcodes following the manufacturer’s protocol (*Nanopore kit SQK-LSK110/SQK-LSK114 and EXP-PBC096*)

### Sequencing

The prepared library was sequenced on a GridION Mk1 platform using MinION R9.4.1/MinION R10.4.1 flowcells and MinKNOW Software with a Guppy basecaller.

### Bioinformatics

#### Pipeline running

We run HERCULES on an HP Z4 workstation equipped with a CPU Intel Xeon W-2235 @ 3.80GHz that provided 12 threads and 128Gb of RAM. Linux Mint 20 was the operating system. Except for the noise cutoff, which was set to 0.1, the rest of the parameters were the default. See Supplementary Methods for a complete description of HERCULES.

#### Statistics

All statistical analyses and plots were done in R^24^.

#### Data normalization

To make wastewater and clinical sampling comparable, wastewater data was normalized using two aggregation steps. In the first aggregation step, all the samples from the same week were used and then in the second aggregation step the ratios of mutations and lineages were weighted according to the population living in each location. The population sizes were obtained from the Norwegian Statistical Center (https://www.ssb.no/).

#### Generation of HERCULES-ready sequences from FASTA files

To use HERCULES to analyze clinical data, nucleotide sequences in FASTA format were aligned to the Wuhan-Hu-1 reference using Nextclade^25^. The aligned sequences were imported into R^24^ using the SeqinR package^26^. Next, the aligned sequences were trimmed to contain just the region of interest. The sequences were saved as a TSV file with two columns, one column containing the trimmed sequences and the other in the same region from the Wuhan-Hu-1 reference. The collection dates of the samples were encoded in the name of the files (e.g., GISAID.01.01.2024_EPISL14259917). This format is fully compatible with HERCULES, and it is the format in which the database containing previous samples is stored.

### Data availability

The 214 FASTQ files used in this paper are deposited in ENA under the project PRJEB76651 and the 8087 sequences of the clinical data used to evaluate HERCULES can be obtained from GISAID using the EPI_SET ID: EPI_SET_240806fr.

### Code availability

HERCULES source code and HERCULES Docker image are freely available on GitHub (https://github.com/garcia-nacho/HERCULES).

## Ethical considerations

This study is part of the project *“Testing an implementation of wastewater-based surveillance of SARS-CoV-2 and its variants”* which was approved by the Regional Committee for medical and healthcare research ethics-South-East C region (Reference REK 454077) based on the Norwegian Health Research Act.

Wastewater surveillance data are derived from pooled environmental samples and not from individuals, therefore informed consent is not applicable.

## Autor contribution

KB, LM, and IG conceptualized the paper. IG designed and implemented HERCULES. RR sequenced the wastewater samples. AR analyzed and evaluated the Pangolin lineage assignment. EHM planned and managed the wastewater pilot, including facilitation of data collection, data preparation and helped in data interpretation. IG drafted the manuscript. IG, LM, RR and KB wrote the manuscript. All authors revised the paper.

## Data Availability

All data produced is deposited online at the European Nucleotide Archive under the project PRJEB76651. The source code and Docker images are available on GitHub (https://github.com/garcia-nacho/HERCULES)

https://www.ebi.ac.uk/ena/browser/view/PRJEB76651

https://github.com/garcia-nacho/HERCULES

## Acknowledgments

We would like to thank Amanda G. Qvesel and Lasse D. Rasmussen from the SSI in Denmark for helpful discussions. We thank the municipal doctors and staff at the wastewater treatment plants for their willingness to participate in the project and providing samples. We thank key personnel at the contract laboratory Nemko Norlab for the preparation of samples. The authors also thank the EU-WISH (EU-Wastewater Integrated Surveillance for Public Health) team at NIPH for their helpful input in the evaluation of the project. We appreciate the highly skilled technicians at the Influenza and other respiratory viruses section at the NIPH involved in the project, especially Marie Madsen Paulsen and Malene Knutsen and we would also like to thank other colleagues, especially Jon Bråte, at the virology department for fruitful and helpful discussions.

---

**Figure S1:**
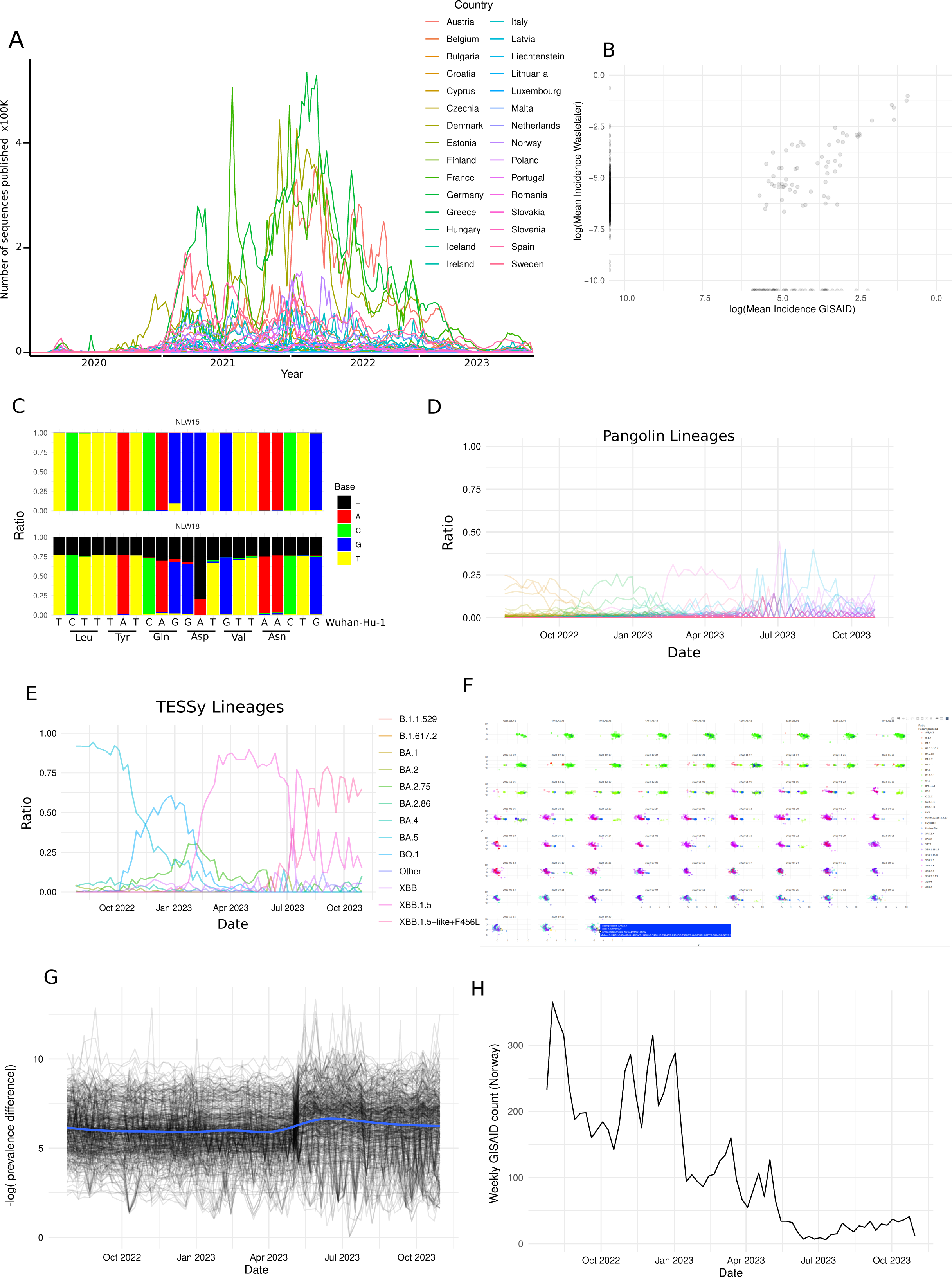
Data controls. **A.** Line plot showing the number of sequences from clinical samples deposited in GISAID by the different European countries. The countries are represented with different colors. **B.** Scatter plot showing the correlation between the average incidence of the *mutation-lineages* found in clinical samples and wastewater. Each point represents a combination of mutations (i.e., a *mutation-lineage*). The points aligned on the bottom or left borders represent *mutation-lineages* not found on wastewater and GISAID respectively. **C.** Bar plots showing the proportion of the different nucleotides mapped on the 614^th^ codon area for NWL15 and NWL18 lineages. The nucleotide and amino acid sequences of the Wuhan-Hu-1 reference are shown. The different colors show the different nucleotides (black shows that a deletion was mapped). **D.** Line plot representing the temporal incidence of the different Pangolin lineages in Norway. Each color represents a different lineage. **E.** Line plot representing the temporal incidence of the different TESSy lineages in Norway. Each color represents a different lineage. **F.** Screenshot of the 2D-map HTML widget generated by HERCULES. **G.** Line plot showing the variability in the absolute differences between the detected point mutations (See Fig. 4B). Each line represents a point mutation. The blue line represents the smoothed average of the lines. **H.** Line plot showing the number of weekly Norwegian clinical samples deposited in GISAID.

**Figure S2:**
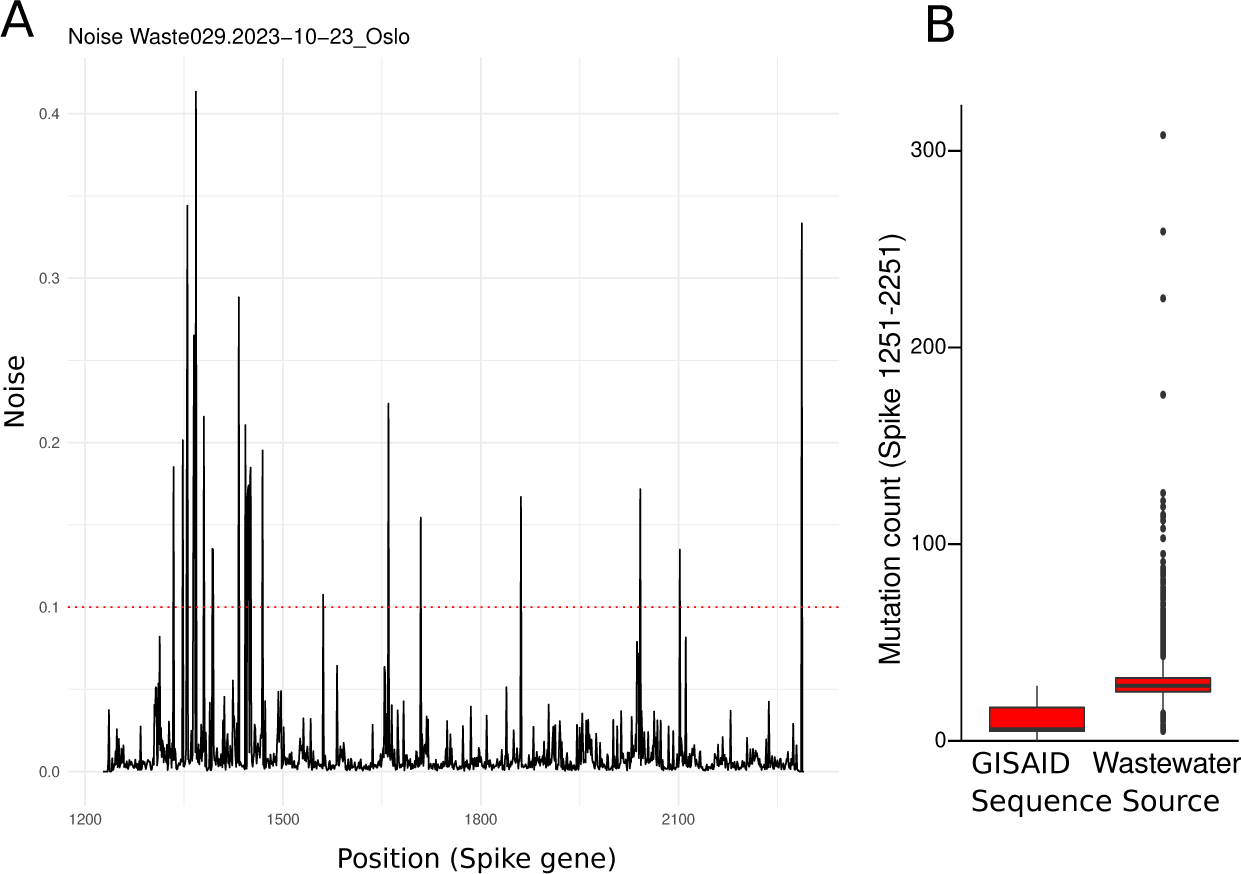
Controls used during HERCULES development. **A.** Representative noise plot generated by HERCULES at running time. On the X-axis the nucleotide position regarding the Spike gene is shown. On the Y-axis the noise (i.e., the ratio of reads that have a nucleotide different from the most common nucleotide found in a particular position). The dashed red line represents the cutoff used by HERCULES to find positions of interest. **B.** Box plots showing the number of mutations at nucleotide level between the positions 1251 and 2251 of the Spike gene in clinical samples (GISAID) and wastewater.

**Table S1:** Average prevalence of the amino acid substitutions identified in wastewater samples and their prevalence in clinical samples in Norway and worldwide.

**Table S2:** Pangolin lineages included in each TESSy lineage by the 8th of August 2024.

