## Supplementary Methods for "Unsupervised detection of SARS-CoV-2 mutations and lineages in Norwegian wastewater samples using long-read sequencing"

**Supplementary Methods:  HERCULES detailed description**

Index:

1. HERCULES implementation details

2. Folder structure

3. Preprocessing and filtering

4. Read mapping

5. Unsupervised identification of positions of interest

6. Sequence alignment extraction

7. Wastewater database

8. Sequence dereplication

9. HERCULES' sequence database

10. Using a set of sequences as a database

11. Lineage map generation

12. Pangolin Lineage classification

13. Pangolin compression

14. Minimum spanning tree calculation

15. Mutation identification

16. Mutation-lineage identification

17. Kmer quantification

18. Sankey plot generation

19. HTML widgets

20. HERCULES' parameters

21. References

### 1. HERCULES implementation details

HECULES is implemented as a Docker image[1] that contains everything needed to run (i.e., scripts, binaries, libraries and databases). The source code and the Docker images are available as a GitHub repository (https://github.com/garcia-nacho/HERCULES).

To improve the efficiency of the pipeline, the most computationally heavy tasks (e.g., mapping to reference, extraction of mapped reads, masking of non-informative positions) are parallelized through the maximum number of available cores minus two. Unfortunately, the larger the number of samples, the more memory HERCULES requires to run. To allow running HERCULES in low-RAM machines, the pipeline can run with four different memory settings (high, medium, low or very low) which split the samples into batches of different sizes (larger for high memory settings). Running the pipeline in lower memory settings will take more time to be completed at the cost of using less RAM.

For reference, the entire Norwegian wastewater dataset can run flawlessly on a computer with 12 threads and 128GB of RAM using high-memory settings.  The parallelization is performed at the minimap tool level (See “Read mapping” section) and in the *ParallelBamExtraction.R* script which uses the *doParallel*, *parallel*, *foreach* and *doSNOW* R packages.

### 2. Folder structure

Before running HERCULES, the samples must be organized into subfolders with all the reads from the same sample stored in the same folder. HERCULES uses the names of the folders as sample names and each sample name has three parameters encoded: Experiment, location and date (e.g., The folder name *RUN01.2024-01-01_Location01* would include the sample taken *at Location01* on the 1st of January of 2024 and it was run in the experiment *RUN01*). To simplify the renaming process, HERCULES comes with a script to rename the samples according to a provided Excel spreadsheet (See https://github.com/garcia-nacho/HERCULES/Template.xlsx for a template of the spreadsheet). If the spreadsheet is present in the main folder, HERCULES will rename the subfolders according to the information provided on it.

#### 3. Preprocessing and filtering

Before processing the samples, all the FASTQ.GZ files are merged into a single file in a folder-wise manner. Low-quality reads are then filtered out. It is possible to use different Q30 quality cutoffs when running HERCULES. The default Q30 cutoff value is 15.

Reads too short (i.e., size below 500nt) or too large (i.e., size above 1500nt) are also filtered out by default. It is possible to change or remove these default values when running HERCULES. All filtering steps are performed using *seqkit*[2].

Although we did not run any primer trimming step in this paper, we implemented an optional trimming step using *cutadapt*[3].

#### 4. Read mapping

Filtered reads are then mapped against the Wuhan-Hu-1 Spike gene using *minimap2*[4] (Li, 2018). *Samtools* (Danecek et al., 2021) is used to filter out supplementary/secondary alignments, optical duplicates and unmapped reads. Next, SAM files are converted into BAM files which are sorted, indexed and piled-up using *samtools*[5].

#### 5. Unsupervised identification of positions of interest

To find the sites that contain mutations, HERCULES runs *NoiseExtractor* (https://github.com/garcia-nacho/NoisExtractor)[6] on the BAM files. Then, HERCULES filters out the sites in which the noise values (i.e., the ratio of reads that do not agree with the nucleotide present in the majority of the reads) are consistent with sequencing errors. HERCULES generates noise plots to help the user to eventually adjust the noise cutoff (Fig. S2A). To perform the filtering step, HERCULES averages the noise and depth of each nucleotide position for all the samples, including the ones present on the HERCULES-Wastewater database. Then, the positions with noises over the noise cutoff parameter (0.1 by default) and an average depth over 20 reads are pre-selected. If the number of pre-selected sites is higher than 50, only the top 50 sites (i.e., those with higher noise) are used further in the analysis. This filtering step prevents the inclusion of too many, most likely false-positive, lineages. To include extra mutations of interest, HERCULES uses an internal or external database of sequences (See below) and includes the positions that differ in at least 5% of the sequences present in the database.

Additionally, HERCULES always analyzes the position 23403 (According to Wuhan-Hu-1) which is where the mutation S:D614G is encoded (A23403G). This serves as a positive control since most of SARS-CoV-2 lineages are expected to carry the S:D614G mutation[7]. Moreover, we implemented a way to supply additional sites of interest to HERCULES. When those additional sites are included, the pipeline includes them in the analysis disregarding their noise.

Sequences generated with long-read sequencing technologies have a relatively high error rate, which means that on average every read will contain several sequencing errors. Indeed, we observed that the number of mutations (i.e., sites that are different from the Wuhan-Hu-1 reference) is much higher in wastewater-derived sequences than in clinical samples (Fig. S2B). The incorporation of positions that do not contain actual mutations in the virus would result in the inclusion of false-positive lineages. Therefore, there exists a strong tradeoff between sensitivity (i.e., HECULES' ability to find a true lineage in the sample) and specificity (i.e., HERCULES’ ability to ignore a false-positive lineage derived from sequencing errors). Although we have observed that a noise cutoff between 0.10 and 0.15 works well, other cutoffs might work better in other settings and testing different noise cutoffs is highly recommended.

#### 6. Sequence alignment extraction

Since raw reads might contain insertions and deletions due to real mutations and/or sequencing errors, they need to be aligned to the reference. This alignment is performed by the mapping tool (See above) and the extraction of the aligned reads in an R-friendly format is done by *bam2msa* (https://github.com/orangeSi/bam2msa). *bam2msa* takes the sorted and piled up BAM files as input and outputs a tabular file containing the reads aligned to the reference.

Nucleotide insertions are a very common sequencing error in ONT (on average we found ~ 4.9 nucleotide insertions on the 1001bp fragment). Moreover, insertions are difficult to handle computationally since they produce displacements in the index of the nucleotides with regard to the reference. HERCULES, therefore, trims out the insertions from the aligned reads.

Finally, HERCULES saves a compressed copy of the table containing the aligned reads and the reference. The sequences in this format are saved by HERCULES and they can be used as input to reanalyze the data when new samples arrive (See "Wastewater database" section).

#### 7. Wastewater database

HERCULES is designed to be a surveillance tool that can reanalyze previous samples and update previous results as new samples are included. To spare time during the analysis, previous HERCULES data are stored in a wastewater database that contains all the information needed to incorporate previous samples into the current analysis. The database has two files per sample, one file containing the noise information of the sample (See "*Unsupervised identification of* *positions of interest*” section above) and a compressed tabular file (See “*Sequence alignment extraction”* section) and HERCULES saves a copy of this database in the Results/WWDB folder.

When running HERCULES it is possible to provide the path to a wastewater database so that the pipeline can integrate previous samples.

Once HERCULES finishes the analysis that contains old and new samples, it rebuilds and stores the entire database in the Results/WWDB folder.

### 8. Sequence dereplication

After removing the non-informative positions from the aligned sequences, the remaining sites are used to aggregate the reads. Each combination of mutations is aggregated in a location and date-wise manner.

### 9. HERCULES’ sequence database generation

To assign Pangolin lineages to the different reads, to compute additional positions of interest, and to compute a lineage-probability table (See “Lineage map generation” section), HERCULES uses an internal database. Although HERCULES has already an internal database embedded in the Docker image, it is possible to use a different set of sequences to be used as the database at running time.

The internal database included in HERCULES’ Docker image was generated using 9210 sequences downloaded from NCBI. To get a representative overview of the different lineages detected up to the 28 of August 2023, we used *covSampler*[8]. Additionally, we added 11 extra BA.2.86 sequences downloaded from GISAID[9] (Khare et al., 2021) to the database.

These sequences were aligned to the Wuhan-Hu-1 reference and assigned to Pangolin lineages using Nextclade[10]. Then, The Spike gene was extracted from the aligned sequences and connected with their Pangolin lineages that were encoded in the sequences' names (e.g., BA.1_SQXXX.fasta). The files with identical sequences and identical lineages were removed and the sequences that were identical at the nucleotide level but that belonged to different lineages were assigned to the closest possible Pangolin lineage (See "Pangolin compression" section).

The sequences were stored in a multifasta file and used as input for the database generation script (i.e., *ProbMatrixGeneratorHercules.R*) which further generates the probability table (See below), the positions of interest, and a HERCULES-ready file to be analyzed alongside the samples.

It is possible to use a different set of sequences as a reference by mapping a folder containing them into the /Reference folder in the Docker's file system (See "Running parameters" section). The different Pangolin lineages must be encoded inside the multifasta file as sequences' names. Lineages must be separated from a sequence index (i.e., a unique number) with an underscore (e.g., BA.2.86_SEQ1). Although HERCULES expects Pangolin nomenclatures, any other lineage classification method can be used. Due to the unsupervised nature of HERCULES, updating the database is not required for *mutation-lineage* mapping and identification; and only becomes relevant to predict Pangolin lineages.

### 11. Lineage map generation

To help the visualization of all the *mutation-lineages* present in the sample, HERCULES generates scatter plots including information about the time and location of the sample.

First, a probability table is precomputed. In this table, all unique Pangolin lineages present in the database are stored as columns, and the sequences of the region of interest stored as rows. Each nucleotide position has 5 rows assigned, one for each possible base plus a row for deletions and in each cell the probability of the presence of a mutation on a lineage was stored. Formally:

$M_{ps,l} = S(L_l \mid ps)$

$$P(L_l|ps|) = \frac{|L_l(ps)|}{|L_l|}$$

Where *M* is a probability matrix; *ps* is the combination of nucleotide position *n* from the set N={*1,…,1001*}; and substitution *s* from the set S={*A,T,C,G,-*}; *l* is a lineage of the set L={$lineage_1$, …. , $lineage_k$} and $P(L_l|ps)$ is the probability of the combination *ps* for the lineage *l*.

Next, a *lineage-mutation* matrix (*C*) is computed for all the aggregated lineages in the sample. C is generated as a binary matrix where the rows are the aggregated lineages (See above) and the columns are all possible nucleotide-substitution combinations. The matrix is filled with zeros and ones according to the absence or presence of a particular substitution in a particular site (i.e., one-hot encoding format).

Then, a *lineage-score* matrix is generated by multiplying the matrices *C* and *M*. In this l matrix, each read gets a score for each lineage based on the mutations it carries. The scores are the sums of the probabilities associated with the lineages for each mutation.

To visualize the lineages in 2D, a dimensional reduction step is performed on the C l*ineage-score* matrix using UMAP[11]. Moreover, HERCULES saves the UMAP coordinates of each single lineage as well as PDF and HTML versions of the plots to facilitate the visual inspection of all the lineages of the sample.

**12. Pangolin lineage classification**

To predict the Pangolin lineage associated with each unique *mutation-lineage*, HERCULES uses its internal sequence database. For each *mutation-lineage* a Pangolin score is assigned based on mutation-profile similarities. The mutations shared by a *mutation-lineage* and a sequence in the database are scored as +1 and the divergent mutations (i.e., the mutations present in the *mutation-lineages* but not in the sequences in the database and vice versa) are scored as -1. The individual scores then are added up and the Pangolin lineage associated with the sequence with the highest score is assigned to the *mutation-lineage*. If several Pangolin lineages are assigned to the same *mutation-lineage*, the closest common Pangolin lineage is assigned to the sequence (See “Pangolin compression” section)

**13. Pangolin compression**

When two or more different Pangolin lineages are equally likely during the Pangolin prediction, HERCULES finds and assigns the closer common Pangolin lineage.

Pangolin nomenclature was designed to carry some phylogenetic information which can be used to find the closest common lineage of several lineages. For example, the closest common lineage to B.1.1.1 and B.1.1.2 is B.1.1.X.

However, due to the increasing number of lineages, the length of the Pangolin names became very long and non-practical. To mitigate this, aliases were created for some relevant lineages. (e.g., JV is the alias for the B.1.1.529.2.75.3.4.1.1.1.1.1.1.7.1.2 Pangolin lineage). When finding the common Pangolin lineages, HERCULES first *unaliases* the lineages using the list of aliases stored in the GitHub repository *cov-lineages/pango-designation*, then it finds the closest common lineage and then it recompresses the closest common lineage to an alias when possible.

HERCULES stores a snapshot of the list of aliases (date of the snapshot Nov 24, 2023) so that HERCULES can run in air-gapped environments.

### 14. Minimum spanning tree calculation

To help understanding the phylogenetic evolution of the virus within the samples, HERCULES generates minimum spanning trees (MST) of the *mutation-lineages*.

MSTs are generated using a modified version of *mst* function from the *ape* R-package[12,13] (Paradis and Schliep, 2018; Garcia et al., 2024).

HTML widgets to visualize the entire MST of all the mutation lineages with incidence over 1% are stored.

### 15. Mutation identification

After aggregating the nucleotide sequences of the positions of interest, the amino acids corresponding to each combination of mutations are obtained. Next, the frequencies of each amino acid substitution are calculated. Finally, HERCULES generates a table and a bar plot per sample to visualize the ratio of the different amino acid substitutions.

### 16. Mutation-lineage identification

HERCULES finds the different *mutation-lineages* by aggregating the combinations of mutations at the amino acid level that are found on the same reads.

Although HERCULES generates a raw table including all *mutation-lineages* present in the sample, it flags the *mutation-lineages* present in at least 10% of the samples with a ratio over 4% in at least one sample. These high-frequency *mutation-lineages* are plotted out for a quick visual inspection.

### 17. *Kmer* quantification

To increase the sensitivity of HERCULES in detecting very low-frequency lineages (i.e., lineages with frequencies below ONT's error threshold), HERCULES is equipped with a *kmer* search tool implemented in R.  The *kmer* search is performed on the aligned and extracted reads including the samples present in the wastewater database.

If one or several *kmers* are provided as a parameter at running time, the reads containing the *kmers* provided will be identified and quantified. Plots to visualize the evolution of the *kmers*' frequencies over time are saved and FASTA files containing the reads carrying the *kmers* of interest are saved.

*Kmers* can be provided as nucleotide sequences (e.g., TAAGCATAGTG), as point mutations alone (e.g., C23271T) or as combinations of point mutations (e.g., C23271T-C23423T-G23222A).

### 18. Sankey plot generation

HERCULES presents information about the *mutation-lineages* and their frequencies as Sankey plots which can be helpful to understand the evolution of the virus. Sankey plots are generated in R using the ggsankey package (https://github.com/davidsjoberg/ggsankey). HERCULES saves one plot per sample.

### 19. HTML widgets

HERCULES generates several HTML widgets that can be visualized in the browser or integrated into surveillance dashboards. HERCULES uses *htmlwidgets* and *trelliscopejs* R packages to generate and save the widgets.

### 20. HERCULES' parameters

HERCULES can take different parameters which are passed through the Docker run command.

The list of parameters is:

-v InputFolder:/Data: Maps the input folder containing the FASTQ files into the /Data folder inside the Docker containers which is where the pipeline script expects them. This parameter is mandatory for HERCULES to run.

-v Database:/Previous: Maps the wastewater database into the /Previous folder inside the Docker container. This parameter is optional, if provided it will incorporate the database into the analysis.

-v ReferenceSequences:/Reference: Maps a folder containing a set of samples to be used as a reference for the analysis. This parameter is optional, if provided it will override the internal database of HERCULES.

-e qual=15: Defines the Q30 filtering cutoff. The default value is 15.

-e start=1250: Defines where the first nucleotide of the Spike region to be analyzed is located. The default value is 1250 and it must be changed only if other amplification primers are used.

-e end=2250: Defines where the last nucleotide of the Spike region to be analyzed is located. The default value is 2250 and it must be changed only if other amplification primers are used.

-e m=500: Defines the minimum read length. Reads shorter than this value are ignored. The default value is 500.

-e M=1300: Defines the maximum read length. Reads larger than this value are ignored. The default value is 1300.

-e trim=0: Defines the primer sizes to be trimmed at both ends of the reads. The default value is 0 (i.e., no trimming).

-e poi='G22895C': Defines the position/positions of interest according to the Wuhan-Hu-1 reference. They can be provided as nucleotide positions (e.g., 22895) or using the standard mutation nomenclature (e.g., G22895C). If more than one position of interest is provided, they must be separated by a comma. The default value is “0” which is the value to skip this step.

-e kmer='TAAGCATAGTG': Defines the *kmer* or *kmers* to look for during the analysis. The *kmers* can take the format of a small sequence (e.g., TAAGCATAGTG) or a combination of one or several point mutations (e.g., G23048A-C23202A). The default value is “0” which is the value to skip this step.

-e mem='high_mem': Defines the memory setting to be used during the analysis and it has four possible values *high_mem*, *med_mem*, *low_mem,* and vlow_mem. Under high memory settings, HERCULES will use more memory, but it will take less time to run. The parameter defines the batch size to be used during some parallelization steps. The default value is *high_mem*
